# Association between obesity and hospitalization in mild COVID-19 young adult outpatients in Brazil: a prospective cohort study

**DOI:** 10.1101/2021.08.04.21261538

**Authors:** Ivaine Tais Sauthier Sartor, Caroline Nespolo de David, Gabriela Heiden Telo, Gabriela Oliveira Zavaglia, Ingrid Rodrigues Fernandes, Luciane Beatriz Kern, Márcia Polese-Bonatto, Thaís Raupp Azevedo, Amanda Paz Santos, Walquiria Aparecida Ferreira de Almeida, Victor Bertollo Gomes Porto, Fernanda Hammes Varela, Marcelo Comerlato Scotta, Regis Goulart Rosa, Renato T. Stein, COVIDa study group

## Abstract

**Background/Objectives:** The aim of this study was to evaluate the association between obesity and hospitalization in mild COVID-19 adult outpatients in Brazil.

**Subjects/Methods:** Adults with signs and symptoms suggestive of acute SARS-CoV-2 infection who sought two hospitals (one public and one private) emergency department (ED) were prospectively enrolled. Patients with confirmed COVID-19 at inclusion were followed by phone calls at day (D) D7, D14 and D28. Multivariable logistic regression models were employed to explore the association between obesity and other potential predictors for hospitalization.

**Results:** A total of 1,050 participants were screened, 310 were diagnosed with COVID-19 by RT-PCR. Median age was 37.4 (IQR 29.8-45.0) years, and 186 (60.0%) were female. Duration of symptoms was 3.0 (IQR 2.0-5.0) days, and 10.0 (IQR 8.0-12.0) was the median number of symptoms at inclusion. A total of 98 (31.6%) were obese, and 243 (78.4%) had no previous medical conditions. Twenty three participants (23/310, 7.4%) required hospitalization during the period. After adjusting, obesity (BMI≥30.0 kg/m^2^) (OR=2.69, 95%CI 1.63-4.83, P<0.001) and older age (OR=1.05, 95%CI 1.01-1.09, P<0.001), were significantly associated with higher risks of hospitalization.

**Conclusions:** Obesity, followed by aging, was the main factor associated with hospital admission for COVID-19 in a young population in a low-middle income country. Our findings highlighted the need for actions to promote additional protection for obese population, such as vaccination, and to encourage lifestyle changes.

## Introduction

Coronavirus disease 2019 (COVID-19) has brought large numbers of patients to medical attention within a short time span for care of a previously undescribed illness, challenging health services and providers to deliver time-sensitive interventions under difficult circumstances. Since the first cases of COVID-19 were documented in China in late December 2019, the epidemiologic focus has largely been on establishing the severity of disease and outcomes among severe patients^1,2^. Older age and some metabolic disorders such as diabetes were early found to be predictors of hospital admission and mortality in patients with COVID-19^3^.

The COVID-19 pandemic has occurred at a time when the prevalence of obesity is increasing all around the world. Obesity is a well-established risk factor for metabolic non-communicable diseases such as type 2 diabetes and hypertension, highly influencing most major cardiovascular diseases. With the worldwide spread of acute respiratory syndrome coronavirus 2 (SARS-CoV-2), obesity has also emerged as an independent risk factor for severe COVID-19, likely related to the adipose tissue inflammation’s effects on the immune system, besides all the metabolic dysfunction associated^4^. In a recent United States Department of Health and Human Services/Centers for Disease Control and Prevention publication, overweight and obesity were both identified as risk factors for the need of mechanical ventilation related to COVID-19, while only obesity was associated with hospitalization and death in an adult cohort of inpatients^5^. However, as risk estimated for severe COVID-19-associated outcomes were measured mostly among adults who received care at a hospital, the current knowledge about the impact of obesity on COVID-19 hospital admission may possibly be overestimated due to a selection bias, as most studies evaluated only inpatients.

Few studies were developed in mild COVID-19 adult outpatients, especially in low-middle income countries (LMIC). Given the uncertainty regarding outcomes in this low-risk outpatient population, and considering the exponential rise in the prevalence of obesity and its association with severe forms of the disease, this study aimed to evaluate the association between obesity and hospitalization in mild COVID-19 adult outpatients in Brazil. We also aimed to describe the clinical characteristics of COVID-19 outpatients and identify other predictors of hospitalization.

## Materials/Subjects and Methods

### 1. Study design and population

This is a prospective cohort study with data collected in referral hospitals from Southern Brazil, one public (Hospital Restinga e Extremo Sul) and the other from the private health system (Hospital Moinhos de Vento). From 13^th^ May to 14^th^ September 2020 we assessed adult (≥18 years old) outpatients who had a visit in the emergency department (ED) health system for COVID-19 symptoms. Inclusion criteria were presenting at least one sign or symptom suggestive of COVID-19 (cough, fever, or sore throat), within 14 days of onset of symptoms. We then created a cohort of those with confirmed COVID-19, defined as a positive result on qualitative reverse transcription polymerase chain reaction (RT-PCR) for SARS-CoV-2.

### 2. Study procedures

At baseline, clinical, demographic, comorbidities, and signs or symptoms suggestive of COVID-19 data were collected. All participants underwent bilateral nasopharyngeal and oropharyngeal swab collection for SARS-CoV-2 detection via RT-PCR assay. The samples were analyzed in the Molecular Biology Laboratory at Hospital Moinhos de Vento, as described previously^6^. Reported weight and height were used to calculate body mass index (BMI). Participants were followed by phone interviews at 7, 14 and 28 days after seeking care for COVID-19 symptoms (study baseline). Up to ten telephone call attempts were made during a period of three days from the D7, D14 or D28. When necessary, text messages were sent in order to notify that the researchers were trying to contact them. If there was no success in contacting one of the time points, the questions were asked retroactively on the next call. The questions were asked directly to the participant or, in his absence or incapacity, for a person authorized by him at baseline. The participant was considered lost when it was not possible to make any contact in different day periods (including failure in 10 call attempts or time exceeded considering 3-day window) within the 28-day follow-up.

During the follow up interviews, participants were asked about the need of hospital admission, use of supplemental oxygen, admission at intensive care units (ICU), and use of invasive mechanical ventilation. All data were collected by trained researchers and using standardized questionnaires developed for the study in the Research Electronic Data Capture software (REDCap)^7,8^.

The outcome of interest was unplanned hospital admission in the period of 28 days. In order to predict the outcome, demographic and clinical characteristics as age at time of enrollment (years), sex, days of onset of symptoms, racial or ethnic group, active or second hand smoke at home, hospital at inclusion, number of symptoms and number of medical conditions were considered. In order to evaluate obesity as a predictor for the outcome of interest, we used BMI and participants were classified as non-obese (<30.0 kg/m^2^), and obesity (≥30.0 kg/m^2^).

### 3. Statistical analyses

Data normality assumptions were verified using the Shapiro-Wilk test for continuous variables, median values and interquartile ranges (IQR) were calculated, and two-tailed Mann-Whitney-Wilcoxon test was used. Pearson’s Chi-square or Fisher’s exact tests were used to evaluate proportions. Uni and multivariable logistic regression analyses were performed to assess the predictors of hospitalization considering the follow up period of 28 days after having a COVID-19 health care visit (study baseline). The association between obesity and hospitalization was assessed using multiple logistic regression models with adjustment for relevant covariates (hospital at inclusion; years of education; number of medical conditions (none, one, two or more); days of onset of symptoms to inclusion; number of symptoms (≤8, 9-10, 11-12, ≥13); and cardiovascular diseases, which includes: ischemic heart disease, heart failure and hypertension). We performed additional models including covariates considered risk factors to hospitalization in order to assess consistency and risk of bias. Odds ratios (ORs) with 95% confidence intervals (CIs) were calculated. All data preprocessing and analyses were performed in R 3.5.0 statistical software^9^.

### 4. Ethics

The study was performed in accordance with the Decree 466/12^10^ of the National Health Council and Good Clinical Practice Guidelines, after approval by the Institutional Review Board (IRB nº 4.637.933). All participants included in this study provided written informed consent.

## Results

In this study 1,050 subjects were screened and 13 were excluded (9 for not meeting inclusion criteria, 2 for not consenting, and 1 withdrew consent, and 1 without SARS-CoV-2 RT-PCR result), and 727 participants with negative RT-PCR results. A total of 310 adults with confirmed SARS-CoV-2 were included. The success in the 28-days follow-up was obtained for 297 (95.8%) participants, as shown in Figure 1.

**Figure 1.**
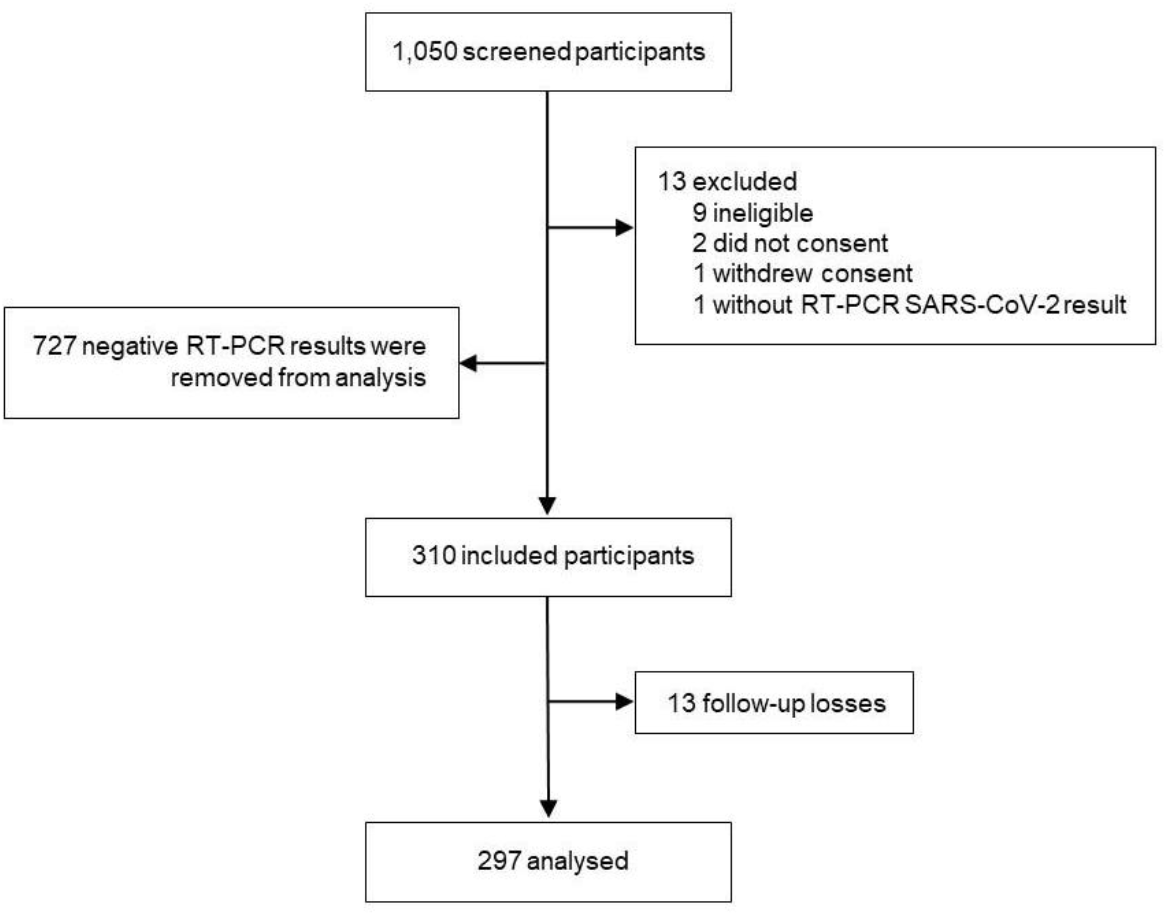
Study flowchart. Thirty three participants out of 297 answered the questions retroactively since there was no success in contact at that time point. Twelve responsible family members were the respondents. (RT-PCR) Reverse transcription polymerase chain reaction. (SARS-CoV-2) Severe acute respiratory syndrome coronavirus 2.

At baseline, the median participants’ age was 37.4 years (IQR, 29.8-45.0). Out of 310 participants, 186 (60.0%) were female, 222 (71.6%) declared themselves caucasian, and 173 (55.8%) were from the public hospital. The median days of symptoms was 3.0 (IQR, 2.0-5.0), and 10.0 (IQR 8.0-12.0) was the median number of symptoms at inclusion. The most commonly reported symptoms were headache (90.6%), cough (87.4%), malaise (77.1%), and myalgia (76.5%). Few participants were active or second hand smoker at home (17.7%), and more than half was overweight (112, 36.1%) or obese (98, 31.6%). Most participants (78.4%) had no previous medical conditions. Hypertension (12.6%), asthma (6.1%) and type 1 or type 2 diabetes (3.5%) were the most frequent baseline comorbidities reported.

At the 28-day follow-up, 23/310 (7.4%) participants required hospitalization. The median BMI of hospitalized participants was 32.4kg/m^2^ (IQR, 30.2-34.1), whereas for the outpatients was 27.7kg/m^2^ (IQR, 24.3-30.5) (P<0.001). The median of the duration from onset of symptoms at the hospitalizations and at enrollment was 14.0 (IQR, 8.0-17.5) and 9.5 (IQR, 6.0-13.5) days, respectively. During the hospitalization, 15/23 (65.2%) participants required only use of supplemental oxygen, 6/23 (26.1%) were admitted to ICU, 2/23 (8.7%) used invasive mechanical ventilation, and one participant died (1/23, 4.3%).

Participants who required hospitalization were significantly older (P=0.005) and obese (P<0.001) compared with those who did not. No other differences were observed between groups, as shown in Table 1. In multivariable logistic regression analysis age and obesity remained as an important predictor of hospital admission in our cohort (age: OR=1.05, 95%CI 1.01-1.09; obesity: OR=2.69, 95%CI 1.63-4.83). When adjusted for hospital of inclusion, years of education, number of medical conditions, cardiovascular diseases, days of onset of symptoms and number of symptoms at enrollment, age and obesity remained associated with hospitalization, as shown in Figure 2.

**Table 1.**
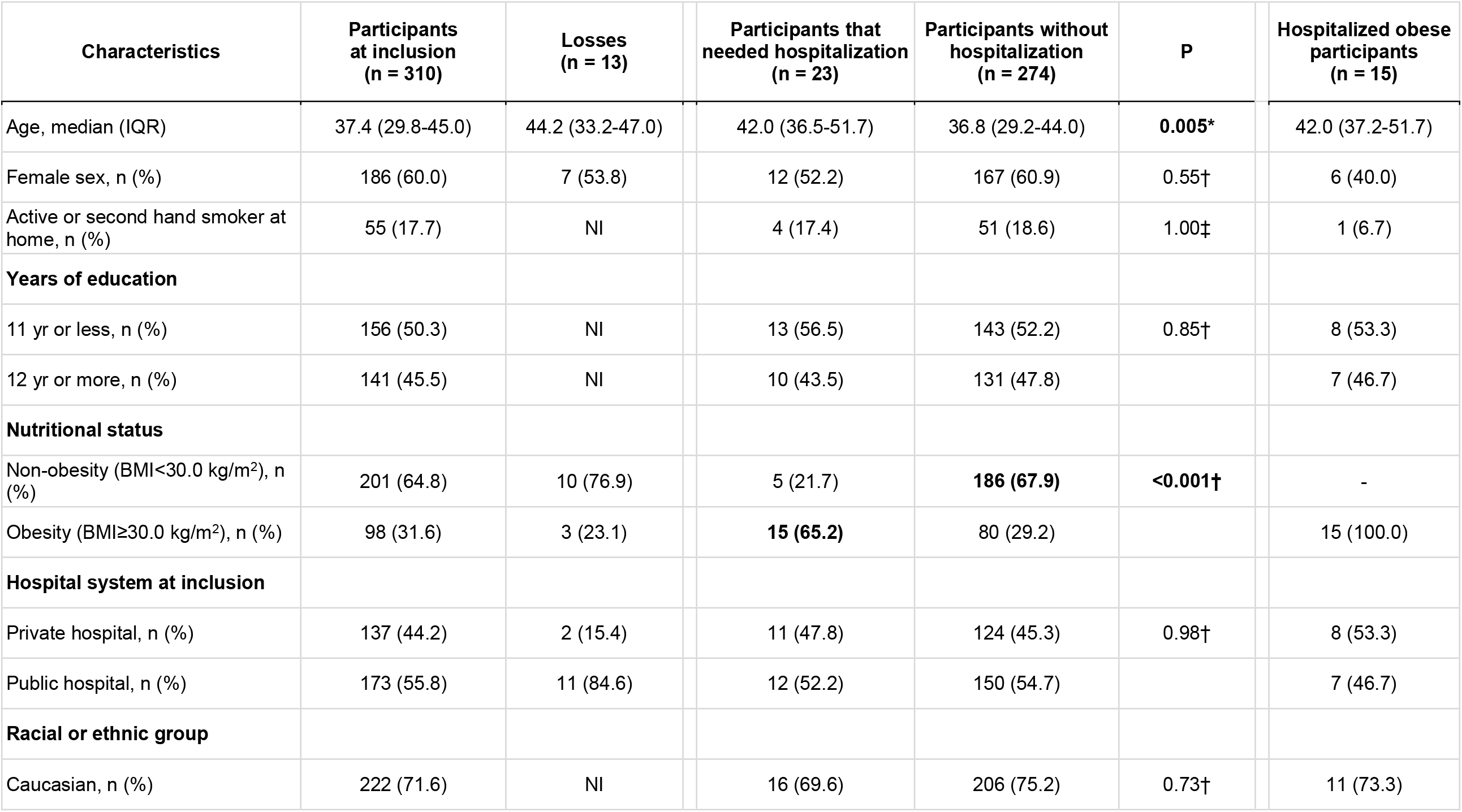
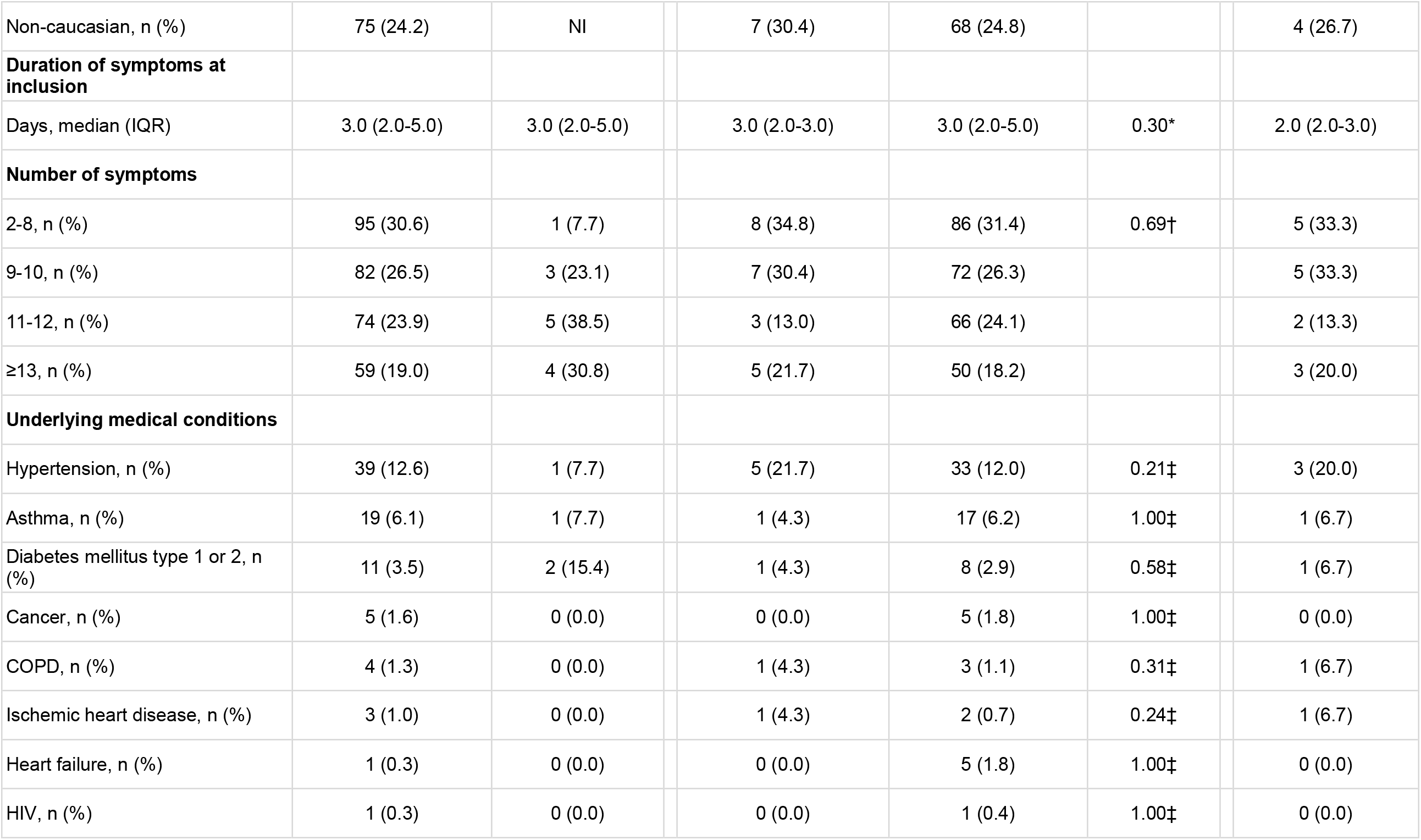
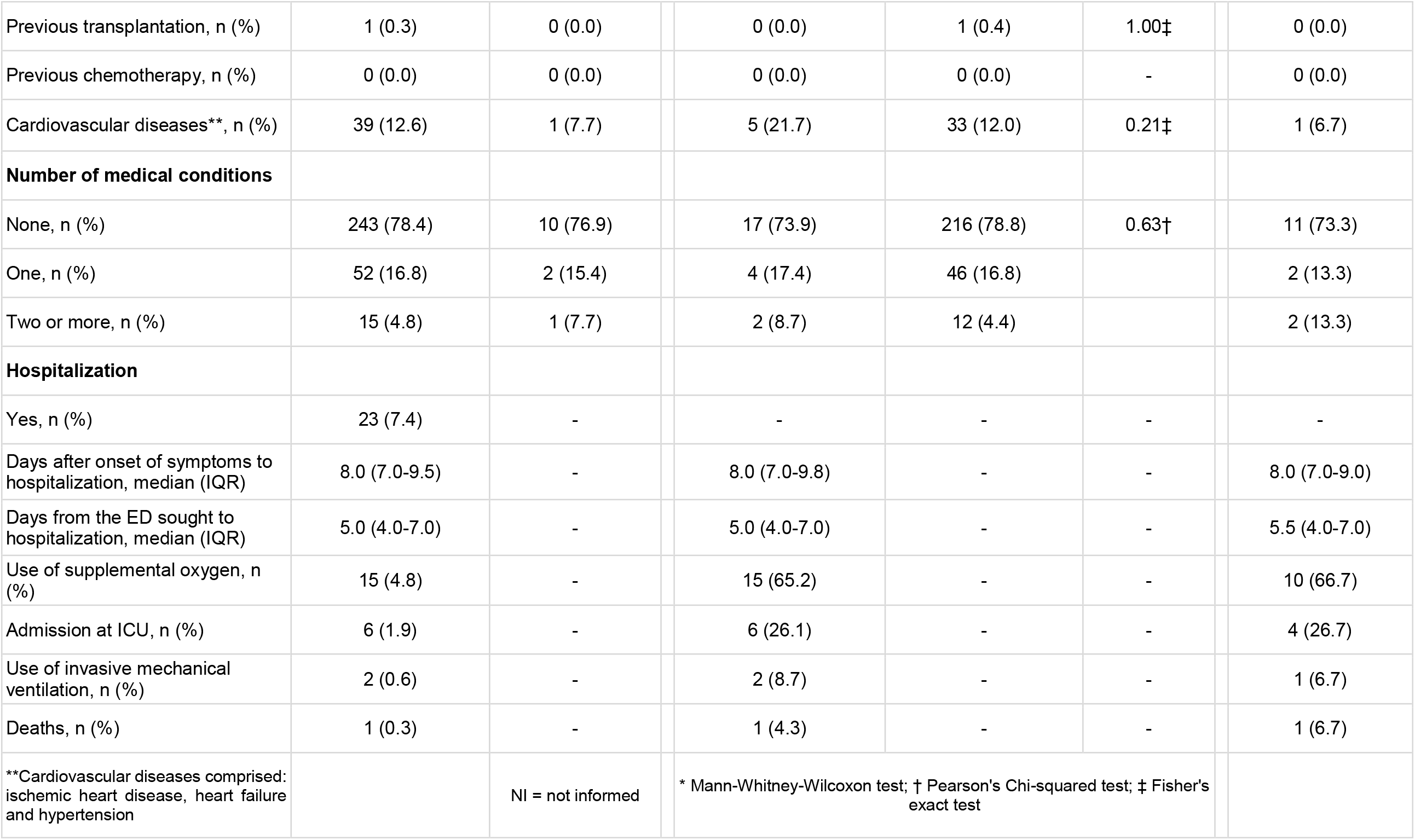
Demographic and clinical characteristics of included subjects.

**Figure 2.**
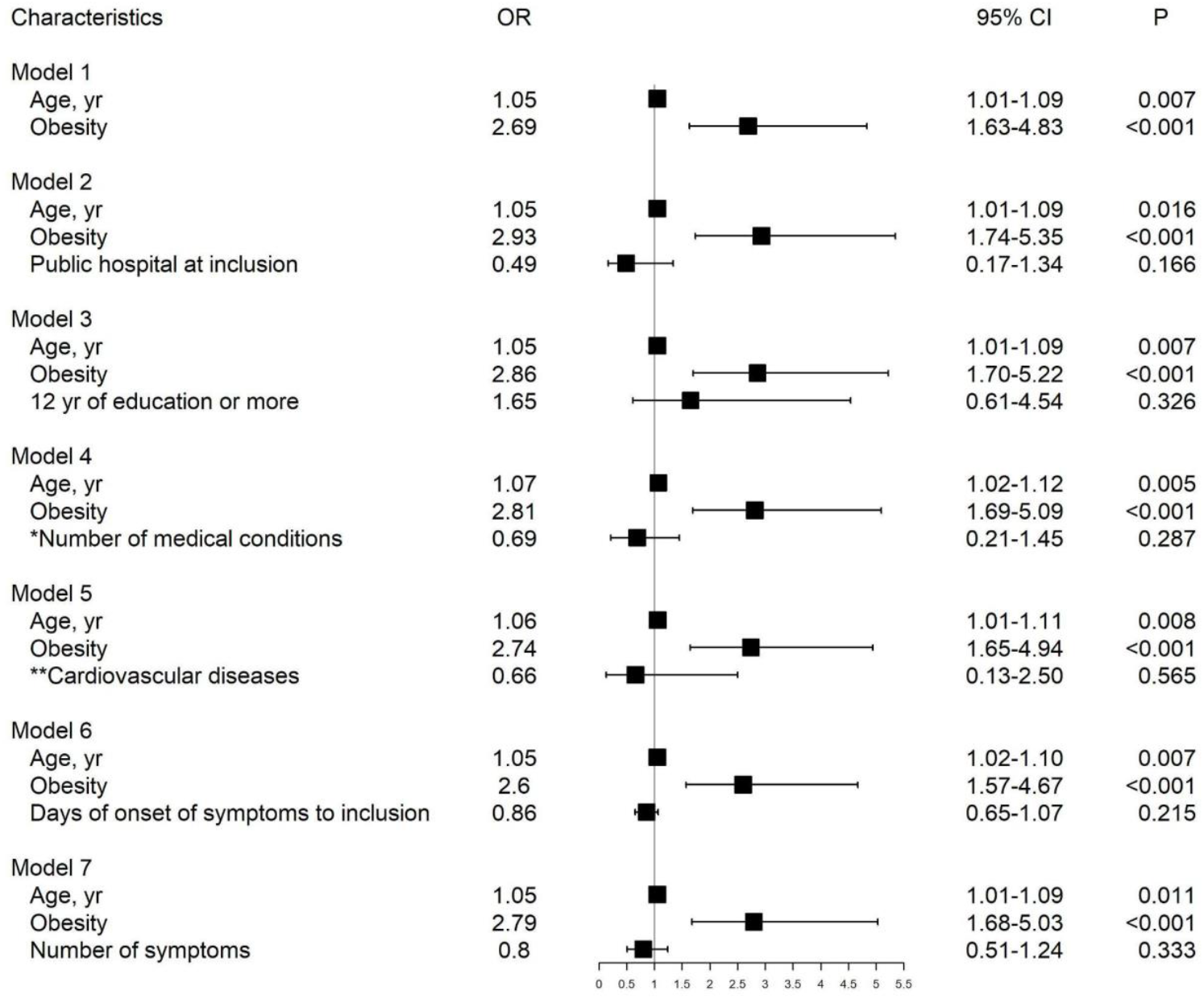
Forestplot of multivariable logistic regression models adjusting age and obesity for the covariates: hospital of inclusion (reference: private hospital at inclusion), years of education (reference: 11 years or less, which corresponds to complete high school education), number of medical conditions (reference: none), cardiovascular diseases (reference: no), days of onset of symptoms (continuous) and number of symptoms (reference: ≤8 symptoms) at enrollment. *Number of medical conditions comprised the sum of cases of: hypertension, asthma, diabetes mellitus type 1 or 2, cancer, chronic obstructive pulmonary disease, ischemic heart disease, heart failure, human immunodeficiency virus and previous transplantation. **Cardiovascular diseases comprised the presence of: ischemic heart disease, heart failure or hypertension.

Among the 15 obese participants with COVID-19 admitted at hospital within 28 days of follow up, the median age was 42.0 years (IQR, 37.2-51.7), 9 (60.0%) were male, 8 (53.3%) were admitted at private hospitals. Four (26.6%) participants reported underlying medical conditions (hypertension, diabetes mellitus, asthma, chronic obstructive pulmonary disease or ischemic heart disease). The median days of onset of symptoms to hospitalization was 8.0 (IQR, 7.0-9.0), and the median days from the ED sought to hospitalization was 5.5 (IQR, 4.0-7.0). Ten (66.7%) participants required supplemental oxygen, 4 (26.7%) were admitted to ICU, one needed invasive mechanical ventilation (6.7%) and one participant died (6.7%).

## Discussion

Obesity is a recognized risk factor associated with worse outcomes related to COVID-19; however, most of the previous studies were carried out in a population of patients already admitted to the hospital or at a high admission rate due to the disease^11^. In this study, we evaluated the risk of hospital admissions in LMIC, through an outpatient cohort of young patients with symptoms of COVID-19 enrolled early in the course of the disease. Our results identified, for a low-risk and young population, that obesity was the main predictor of hospitalization for COVID-19, with approximately 3-fold increase in the risk of hospital admission in comparison to non-obese patients.

The world is still struggling to fight the COVID-19 pandemic. According to the World Health Organization (WHO), the mortality rate from COVID-19 is 0.5-1.0%^12^, while the hospitalization rate for the disease is 5.0-15.0%. Based on currently available information, the Centers for Disease Control and Prevention has identified severe obesity as an important risk factor for worse prognosis and higher mortality in patients with COVID-19, while any degree of obesity has been associated with COVID-19 poor prognosis. In our study, 7.4% of our sample required hospital admission, similarly to data from literature. Most participants were overweight/obese at baseline, but only obesity predicted the main outcome. In a meta-analysis of 75 studies^13^, obesity had a 46.0% higher risk for being COVID-19 positive, a 113.0% higher risk of being hospitalized, and a 48.0% increased mortality. There are many mechanisms that jointly may explain this impact; although the effect of obesity on the immune system remains not completely established, it appears to have an effect independent of coexisting risk factors. Data from literature also identify a dose-response relationship between BMI and COVID-19 worse outcomes^5^.

In addition to metabolic factors that seem to impact the severity of COVID-19, advanced age has been identified as one of the major factors associated with worse outcomes in the pandemic, being one of the most important predictors of hospital admission^11^. In our sample, the mean age of participants was younger than most studies in the literature; however, we also identified an increased risk of hospitalization for COVID-19 with each increased year of age. Concerns brought by our data include the fact that the predictors of higher risk for COVID-19 involve populations especially in need of care by health system^5^. The obese and elderly individuals are possibly the ones that will be mostly compromised by the social distancing that the pandemic generates, possibly resulting in worse health habits, such as physical inactivity and unhealthy eating habits, besides increased emotional distress. This could further aggravate the risk related to COVID-19 for this population, unless governmental and health measures, such as vaccination and strategies to increase consumption of healthier foods, stimulate physical activity and promote mental health^13^, aimed at the health of this population, starts to be prioritized.

Our study has limitations. Our analysis was based on a convenience sample. Moreover, our participants represent a single geographic region of a heterogeneous country, such as Brazil. However, worldwide prevalence of obesity is a public health concern and our findings are in line with previous reports^14–17^. As another point to address, the two centers included in this study represent different populations, public and private, which may characterize different clusters and limit the interpretation of our results for the sample as a whole. Also, most of our data represents self-report and may be subject to bias. Despite that, we believe that our results add to the literature as they represent an outpatient prospective cohort of young individuals with low risk for complications related to COVID-19 in a LMIC, identifying important risk predictors for this population. Moreover, a similar impact of obesity was consistently found despite adjusting for several possible confounders.

In conclusion, our data identified that obesity, in addition to age, was the most important risk factor predicting hospital admission for COVID-19 in a young population in Southern Brazil. These results highlight the importance of health support strategies aimed at this population, in order to promote additional protection, such as vaccination, and to encourage lifestyle changes.

## Data Availability

All data are available in the manuscript.

## Acknowledgments

We thank the Scientific Committee of the Research Support Nucleus (NAP) of Moinhos de Vento Hospital for technical-scientific consultancy. We thank the inclusion personal, laboratory team, and site staff from Hospital Moinhos de Vento and from Hospital Restinga e Extremo Sul. Aline Andrea da Cunha, Joao Ronaldo Mafalda Krauser, Paulo Sergio Kroeff Schmitz, Sidiclei Machado Carvalho, Fabio Jose Rockenbach, Kelly Viegas Antunes, Marcelo da Silva Ferreira, Rafael Garcia Trindade, Thayna Silva Lino, Tiago da Silva Silvano, Adriana da Silva Silveira, Alceu Kuckoski, Ana Paula da Silva Lopes, Andreia Escobar da Costa, Clarice Cardoso Machado, Erica Vieira da Silva, Evelin Inácia da Silva, Luciana Rodrigues Ribeiro, Marcely Mayr da Costa, Morgana Thais Carollo Fernandes, Rafael da Silva Cassafuz.

## COVIDa study group

Adriane Isabel Rohden, Ana Paula dos Santos, Camila Dietrich, Caroline Cabral Robinson, Catia Moreira Guterres, Charles Francisco Ferreira, Débora Vacaro Fogazzi, Denise Arakaki-Sanchez, Fernanda Lutz Tolves, Fernando Rovedder Boita, Francieli Fontana Sutile Tardetti Fantinato, Gisele Alcina Nader Bastos, Jaina da Costa Pereira, Maicon Falavigna, Maristênia Machado Araújo, Patricia Bartholomay Oliveira, Shirlei Villanova Ribeiro, Thainá Dias Luft, Tiago Fazolo.

